# Rethinking Malaria Seasonality: Humidity-Driven Transmission Shifts and Emerging Hotspots in Zambia (2009–2023)

**DOI:** 10.64898/2025.12.05.25341688

**Authors:** Therese Shema Nzayisenga, Nyuma Mbewe, Kelvin Mwangilwa, Jonathan Mwanza, Stephen Bwalya, Ignatius Banda, Cheepa Habeenzeu, Malambo Mutila, Paul Msanzya Zulu, Loveness Nikisi, Freddie Masainga, Allan Mayaba Mwiinde, Peter J. Chipimo, Nathan Kapata

**Affiliations:** Zambia National Public Health Institute, Lusaka, Zambia; World Health Organization, Zambia Country Office, Lusaka, Zambia; National Malaria Elimination Centre, Lusaka, Zambia; Ministry of Health Headquarters, Ndeke House, Lusaka, Zambia; Meteorological Department, Ministry of Green Economy and Environment, Lusaka, Zambia; School of Public Health, University of Zambia, Lusaka, Zambia; Lusaka Apex Medical University, Lusaka, Zambia and Centre for Global Health and Development in Africa, Lusaka, Zambia

**Keywords:** Malaria, Climate variability, Relative humidity, Spatio-temporal modelling, Hotspot analysis, Seasonality, Forecasting, Malaria Vaccines, Climate sensitive disease, Zambia, Sub-Saharan Africa

## Abstract

**Background:** Climate variability is increasingly altering the distribution and seasonality of malaria in Africa, yet evidence to guide climate-resilient control strategies remains limited. Despite having previously been on course for elimination, Zambia has experienced a resurgence of malaria cases alongside intensifying weather events, underscoring the urgency of adapting interventions to shifting transmission dynamics.

**Methods:** An ecological time-series analysis was conducted using 15 years (2009–2023) of district-level malaria surveillance data from the National Malaria Elimination Centre, climate data from the Zambia Meteorological Department, and satellite products. Monthly incidence was correlated with rainfall, temperature, and relative humidity using a structured additive semiparametric Poisson model accounting for spatial and temporal autocorrelation. Hotspots were detected with SATScan, and changes in seasonal transmission were analysed across three five-year periods. Forecast error variance decomposition and Granger-causality tests assessed the direction and strength of climatic variables’ influence on malaria trends.

**Results:** National malaria incidence increased despite intensified control interventions, with a marked geographic shift from historically high-burden provinces (Luapula, Northern, Eastern) toward emerging hotspots in Northwestern, Copperbelt, and Western provinces. The transmission season extended from the traditional January-April peak to December–June, reflecting more extended periods of conducive climatic conditions. Relative humidity was the strongest and most consistent predictor of malaria incidence (p < 0.001), surpassing rainfall and temperature in explanatory power. Predictive models suggest that without enhanced interventions, rising temperatures and humidity will continue to drive increases in incidence through 2030.

**Conclusions:** Malaria transmission in Zambia is becoming more prolonged, spatially dynamic, and increasingly climate sensitive. Integrating real-time climate surveillance, adaptive vector control and synchronised vaccine deployment into malaria programming could strengthen elimination efforts and build resilience in climate-vulnerable settings.

**Author Summary:** Using 15 years of national surveillance and climate data, this study provides new evidence that relative humidity, not rainfall or temperature, is the most consistent and powerful climatic predictor of malaria incidence in Zambia. We show that malaria transmission seasons are lengthening, spatial hotspots are shifting, and climate-sensitive drivers are amplifying risk in previously lower-burden areas. These findings challenge long-standing assumptions guiding malaria control strategies, including the timing of vector control and the rollout of new malaria vaccines. By identifying humidity as a key determinant of malaria risk, this analysis provides actionable insights for climate-resilient programming. It supports integrating real-time climate intelligence into elimination efforts across climate-vulnerable regions.

## Introduction

Climate change is increasingly recognised as one of the greatest threats to global health security, with its effects disproportionately impacting countries that have contributed least to its acceleration (1–3). Its health impacts include increased heat-related mortality, food and water insecurity, altered distribution of infectious diseases, and disruption of health systems (2). A large proportion of climate-related mortality is attributable to increased transmission of climate-sensitive infectious diseases (CSID), including malaria (4).

Malaria remains one of the deadliest infectious diseases worldwide, with an estimated 263 million cases and 597,000 deaths in 2023, an increase of about 11 million cases from 2022. About 94% of global cases and 95% of deaths occurred in sub-Saharan Africa (5). Since 2000, malaria interventions have averted an estimated 2.2 billion cases and 12.7 million deaths. Yet, progress toward the WHO Global Technical Strategy milestones has stalled, and incidence and mortality remain more than double the 2023 targets (5,6).

Despite significant global and national investments, endemic countries remain far from achieving elimination, highlighting the need for climate-resilient strategies that address both biological and environmental drivers of transmission (7–9). Climate-driven changes in precipitation, temperature and humidity can extend periods of environmental suitability for malaria transmission (10,11). High-burden, climate-vulnerable settings are underrepresented in the literature exploring the link between climatic shifts and disease transmission (12).

Zambia, for example, is among 21 African countries that have achieved reductions since 2015 but remains off track for the 2025 goal of a 75% reduction (5). In 2019, malaria incidence reached 340 malaria cases per 1,000 population, and parasite prevalence among children under five years rose from 24% in 2018 to 29% in 2019, signalling a troubling upward trend (13,14), and deviating from the national malaria elimination target of 5 cases per 1,000 population by 2026 (8). These trends reflect overlapping global challenges, including health and humanitarian emergencies, financial crises, and climate change, all of which compound malaria transmission risks and complicate the delivery of core interventions (3,5).

Whereas climate change reflects long-term shifts with occasional catastrophic events such as hurricanes and floods, climate variability captures short- to medium-term departures that more directly influence seasonal malaria risk, providing early warning signals for outbreak detection and response planning (15,16). Earlier modelling studies across Zambia have shown malaria incidence peaks during the rainy season, and is influenced by short-term warming and cooling periods, expanding breeding habitats in previously low-risk areas (14,17). Thus, climate variability can increase transmission risk and strain under-resourced health systems (3,7).

However, there is limited district-level evidence quantifying how specific climate parameters, particularly relative humidity, rainfall and temperature, shape spatial and temporal malaria trends in Zambia. A clearer understanding of these relationships is essential for designing climate-resilient malaria strategies and improving outbreak preparedness. Therefore, this study assessed 15 years of climate and malaria surveillance data to determine how climate variability influences transmission patterns across Zambia and to identify adaptation measures that can be integrated into national malaria control efforts.

## Methodology

### Study Design

This study constitutes the quantitative component of a mixed-methods, cross-sectional study examining the impact of climate variability factors, specifically rainfall patterns, temperature and humidity, on malaria incidence in Zambia. An ecological time-series design was used to examine the relationship between climate variables and malaria trends over a 15-year period (2009-2023). This design enabled both temporal and spatial assessments of climate-sensitive malaria patterns and facilitated the identification of emerging hotspots relevant for climate-resilient planning.

### Study Setting and Context

Zambia is a landlocked country in Southern Africa with a population of approximately 22 million people (18) and ranks among the 20 high-burden countries globally (5). Malaria is endemic and perennial, with varying incidence rates across provinces due to geographic, climatic, and socio-economic differences (19). The country has a tropical climate, with higher rainfall in the north. The rainy season runs from October to April, but in recent years, Zambia has experienced increased flooding associated with El Niño, followed by severe droughts, including the worst drought on record in 2024 (20). These climatic extremes underscore the importance of understanding how short- and long-term variability influence malaria transmission

### Data Sources

#### Malaria Surveillance Data

Malaria incidence and mortality data were sourced from the National Malaria Elimination Centre (NMEC) through the District Health Information System Version 2 (DHIS2). Data included monthly reported total malaria cases (confirmed and clinical), malaria-related deaths and district population estimates for denominator calculations. Data covered the period from January 2009 to December 2023. Incidence rates were calculated using population data from the Zambia Statistical Agency (ZAMSTAT).

#### Climate Data

Climate data were obtained from the Zambia Meteorological Department at 42 manual observation stations across the country, supplemented with satellite-based datasets available online at https://www.ncei.noaa.gov/cdo-web/. Supplementary products included satellite-based rainfall products from the Climate Hazards Group Infra-Red Precipitation (CHIRPS) dataset and Japanese 55-year atmospheric reanalysis (JRA-55) data. Station data were quality-controlled and merged with satellite products using the Climate Data Tool (CDT) to improve spatial coverage for all 116 districts.

Meteorological and satellite datasets were integrated and validated at a 90% confidence level to provide district-level estimates for daily and monthly precipitation (mm), minimum and maximum temperatures (°C), average temperature, and relative humidity (%) were validated using the Climate Data Tool (CDT) and aggregated to monthly and annual averages. A 5-year rolling average was applied to temperature and humidity; rainfall was summed over time.

#### Geospatial Data

District shapefiles were obtained from ZAMSTAT. Due to administrative changes between 2009 and 2023 (from 75 to 116 districts), harmonised shapefile versions were used to ensure accurate temporal mapping and hotspot detection in QGIS.

### Data Management and Processing

The extracted data were checked for completeness and cleaned in RStudio. Climate and malaria datasets were linked by district and month-year. Malaria incidence was calculated as cases per 1,000 population per month. Districts were geo-referenced using centroid coordinates.

### Statistical and Spatial Analysis

#### Trend Smoothing and Hotspot Detection

Geo-spatial visualisations were created by overlaying incidence data on district shapefiles in QGIS. To confirm consistent high-burden areas, administrative districts with persistently high malaria incidence over the 15years were plotted against hotspot analysis output from SATSCAN. Three-year moving averages of malaria incidence were calculated to smooth trends.

#### Climate Trend Analysis

Climate trend analysis was performed using rainfall, temperature, and relative humidity data extracted for each district. Rainfall trends were analysed using the Expert Team on Climate Change Detection and Indices (ETCCDI) methodology via CDT. The Mann-Kendall test, a non-parametric method, was used to detect monotonic trends in environmental. Only indices relevant to rainfall were analysed across all districts, as indicated in Table 1.

**Table 1.** List of Expert Team on Climate Change Detection and Indices (ETCCDI) Extreme Precipitation indices.

| ID | Indicator Name | Definitions | Units |
| --- | --- | --- | --- |
| **CDD** | Consecutive dry days | Maximum number of consecutive days with RR ≥ 1mm | Days |
| **CWD** | Consecutive wet days | Maximum number of consecutive days with RR ≥ 1mm | Days |
| **PRCP**<br>**TOT** | Annual total wet-day precipitation | Annual total PCP in wet days (RR ≥ 1mm) | mm |
| **R20** | Number of very heavy precipitation days | Annual count of days when PCP ≥ 20mm | Days |
| **R95p** | Very wet days | Annual total PCP when RR > 95th percentile | mm |
| **R99p** | Extremely wet days | Annual total PCP when RR > 99th percentile | mm |

PCP = Precipitation
RR = Rainfall

### Estimating Vector Autoregressions (VARs)

Twelve lags were added to each variable in each equation to create a VAR model that included the monthly malaria and climate variables (**Table S1)**. Lag-length selection was guided by the Final Prediction Error (FPE) criterion, which indicated that a 12-month lag sufficiently captured climatic and epidemiological dynamics (21,22). The VAR approach mitigated spurious correlations driven by autocorrelation or shared time trends.

#### Causality Assessment

Granger-causality and instantaneous causality tests were employed to determine whether variations in climate parameters influenced malaria incidence. Forecast Error Variance Decomposition (FEVD) was used to quantify the proportion of malaria variability explained by each climatic factor over time to determine if changes in one variable would influence innovations in another variable, aiding in predicting the other variable a step ahead of schedule (23).

### Structured Additive Regression Modelling

A structured additive semiparametric Poisson regression model (geoadditive model) was applied to assess the association between climate variables and malaria incidence. Independent variables included total monthly rainfall, minimum and maximum temperature, and relative humidity. Models accounted for seasonality, long-term trends, spatial autocorrelation and district-level random effects.

Spearman’s rank correlation coefficients were calculated for three time periods (2009 – 2013, 2014 – 2018, and 2019 – 2022). Correlation categories were mapped in QGIS to identify districts with consistent climatic influence on malaria incidence. Districts with mean correlation coefficients of 0.3 or higher across all three time frames (spanning at least 10 years) were marked in dark colour. Those with correlations in two of the three periods were indicated in lighter colours for each of the four climatic variables. This approach highlighted areas with consistent climatic influences on malaria incidence over time. Districts with sustained correlations >0.3 across 10 years were classified as climate-sensitive hotspots.

### Ethical Considerations

The study used secondary, anonymised routine data from national sources. Ethical approval for this study was obtained from the Lusaka Apex Medical University Institutional Review Board and the National Health Research Authority (Ref No: NHRA000013/23/08/2023). Permission to access data was provided by the Ministry of Health and the Zambia Meteorological Department. Provincial Health Directors were written to inform them of the study, and respective permissions were obtained.

## Results

### Seasonality of Malaria Incidence and Climate Variability

Between January 2009 and December 2022, malaria incidence in Zambia showed a strong and predictable seasonal pattern closely aligned with rainfall and humidity. Fig 1 shows how rainfall exhibited distinct annual peaks that coincided with increases in relative humidity, while malaria incidence consistently increased after these peaks. Temperature showed far less seasonal variability and demonstrated a weaker association with shifts in malaria incidence, remaining comparatively stable throughout the study period. Although the seasonal pattern persisted, the incidence peaks became more pronounced from 2016 onward, with particularly elevated levels between 2021 and 2022, despite continued implementation of malaria control interventions. This decoupling between intervention scale-up and malaria trends underscores the persistent influence of climate on malaria transmission.

**Fig 1:**
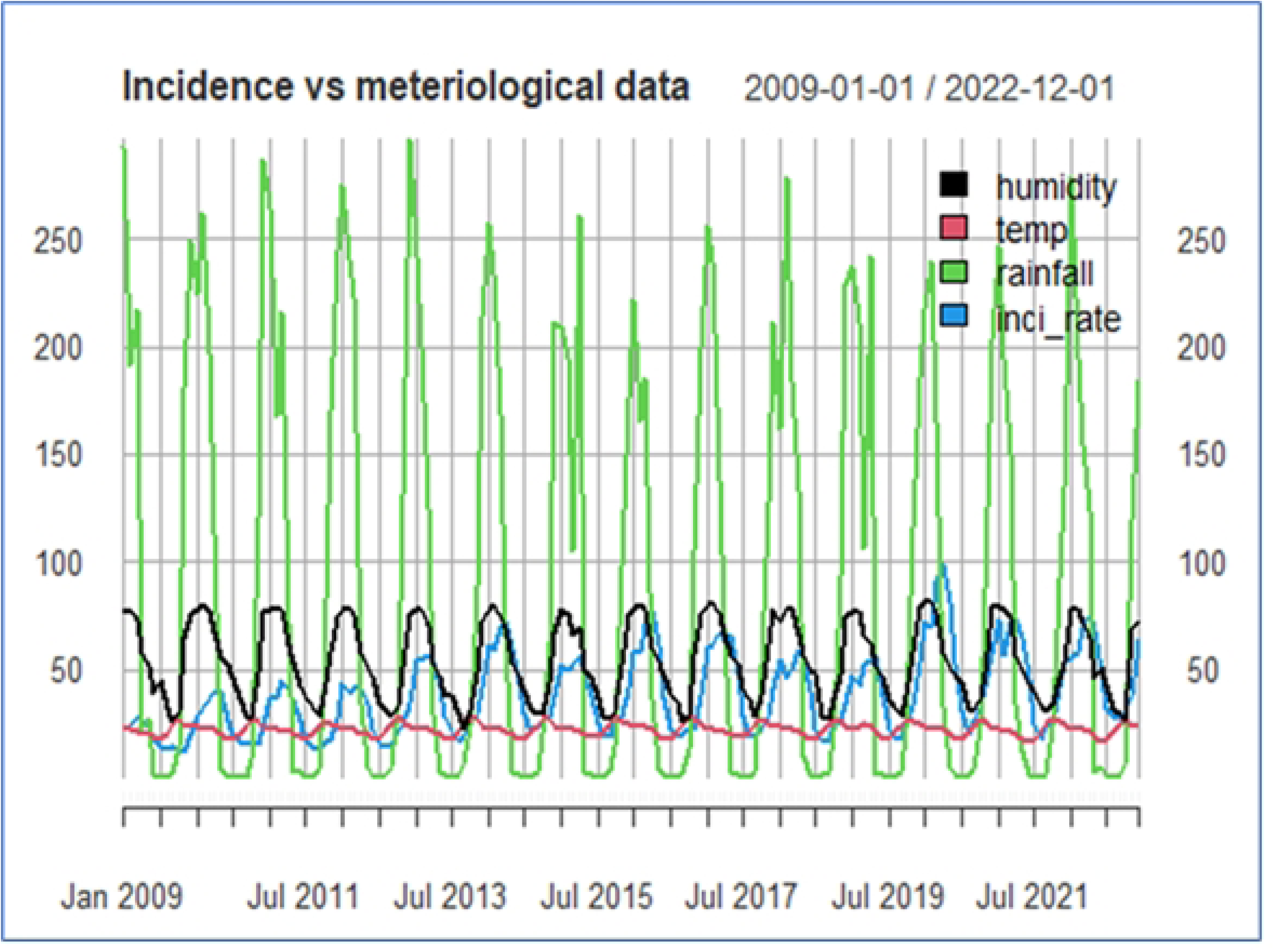
Monthly time series (2009–2022) of malaria incidences per 100,000 population, average temperature (Celsius), average rainfall (mm) and humidity (%) in Zambia. Monthly malaria incidence (blue line) compared with rainfall (green line), relative humidity (black line), and average temperature (red line). Malaria incidence shows strong seasonal peaks corresponding to periods of increased rainfall and humidity, while temperature remains relatively stable across years, indicating consistent climatic influence on transmission over the 15 years.

Cumulative malaria incidence ranged from 185,892 cases in 2016 to 1,326,442 cases in 2020, with a mean annual incidence of 497,694 cases. Across the study period, rainfall ranged from 0.08 mm to 296.62 mm (mean 89.21 mm), temperatures ranged from 16.77 °C to 28.35 °C (mean 22.26 °C), and relative humidity ranged from 22.87% to 81.82% (mean 54.32%). All-time series variables were validated using the Dickey-Fuller (ADF) unit root test (p < 0.05).

Malaria incidence was highest between February and April, and lowest during the dry season (July to October) (Fig 2). Precipitation and humidity peaked from January to March, with median rainfall exceeding 150 mm and humidity exceeding 65%, preceding increases in incidence. By contrast, temperatures fluctuated only modestly, ranging from 20 to 25°C, reinforcing the finding that rainfall and humidity are the primary environmental drivers of malaria seasonality in Zambia.

**Fig 2:**
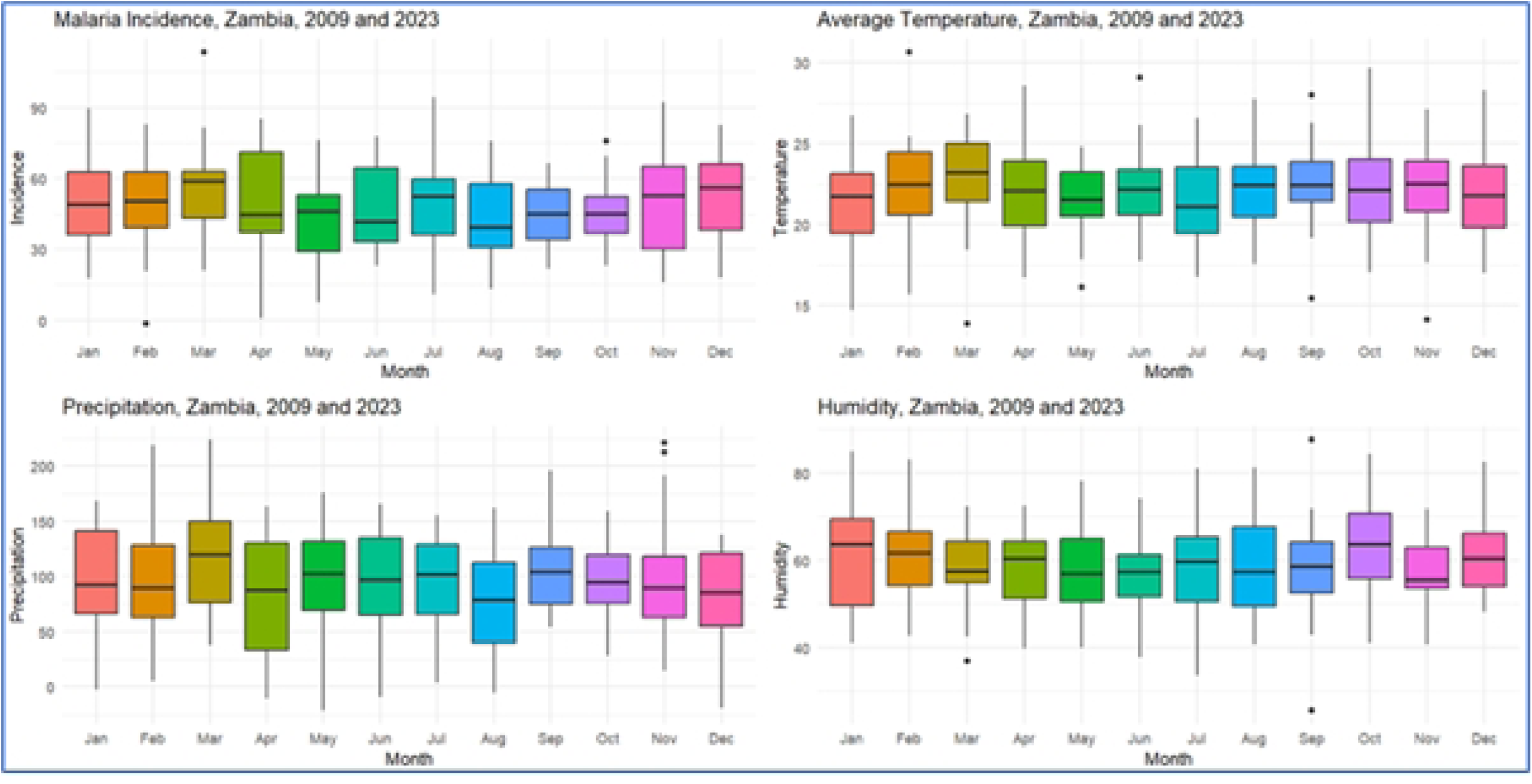
Seasonal distribution of malaria incidence and climate variables in different months from 2009 to 2023 in Zambia. Boxplots display monthly distributions for malaria incidence (top left), average temperature (top right), precipitation (bottom left), and relative humidity (bottom right). Malaria incidence shows pronounced seasonality, with higher median values during the rainy months (March to May).

### Spatial Distribution and Identification of Malaria Hotspots (2009-2023)

District-level maps (Fig 3) revealed persistent malaria hotspots concentrated in northern and north-eastern Zambia, notably Luapula, Muchinga, Northern, and parts of Eastern provinces. In contrast, Lusaka, Southern, and Western Provinces consistently reported lower incidence. Over the 15-year period, high burden districts intensified and expanded geographically, with some districts in Luapula and Muchinga exceeding 1,000 cases per 10,000 population between 2018 and 2023. A notable shift of high-burden zones toward the Northwestern, Copperbelt, and Western provinces emerged after 2020, aligning with regional changes in rainfall and humidity patterns.

**Fig 3:**
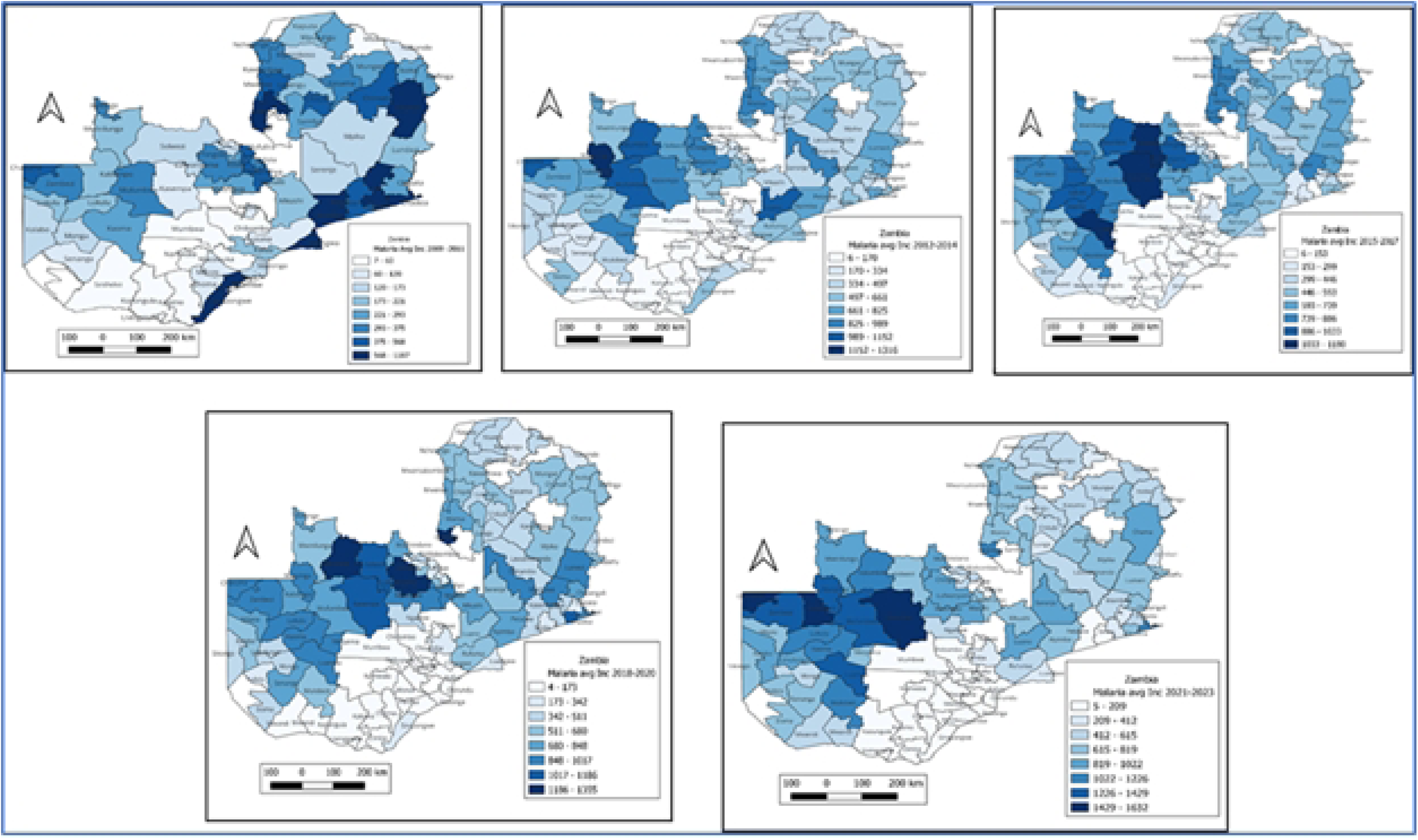
District-level average malaria incidence per 1,000 population across Zambia, 2009– 2023. Maps depict the spatial distribution of malaria incidence averaged over five time periods (2009–2011, 2012–2014, 2015–2017, 2018–2020, 2021–2023). Darker shades indicate higher malaria incidence. Persistent high-burden zones remain in Luapula, Muchinga, Northern, and Eastern provinces, while Southern and Western provinces showed consistently low incidence, emphasising the need for subnational, climate-sensitive interventions.

Nationally, incidence increased from a maximum of 118 cases per 10000 persons during 2009-2011 to 132 per 10,000 cases during 2021-2023. Kasempa District (Northwestern Province) had the highest burden, rising from 505.2 per 1000 in 2017 to 1,317 per 1000 in 2023. Meanwhile, Monze (Southern Province) remained the lowest-burden district, with the incidence increasing from 51.4 per 1000 in 2017 to 101.1 per 1000 in 2023. These spatial patterns closely mirrored rainfall and humidity gradients, with northern regions showing high moisture levels and southern regions becoming increasingly arid.

### Correlation of Climatic Parameters and Malaria Incidence (2009-2023)

#### Rainfall

Rainfall demonstrated significant correlations with malaria incidence in districts with consistently high precipitation, particularly in northern and north-western Zambia (Fig 4a). However, some high-rainfall areas showed weaker associations, suggesting that rainfall alone does not fully determine transmission. Excessive or erratic rainfall may disrupt larval habitats, weakening the strength of the correlation in certain settings.

**Fig 4a:**
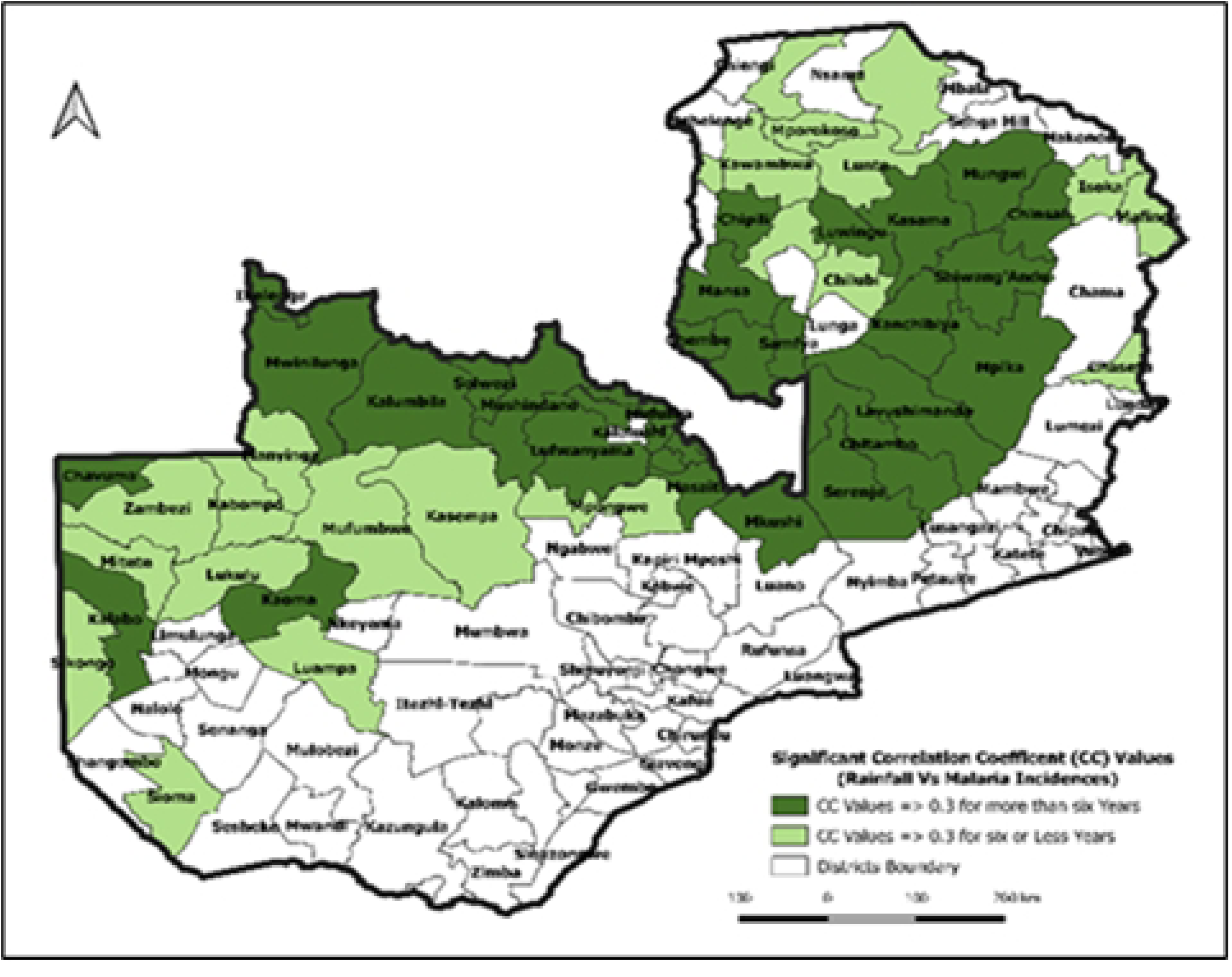
District correlation between rainfall and malaria incidences (2009-2023). Significant correlations were found in 55 districts from 2009 to 2013, 59 districts from 2014 to 2018, and 52 districts from 2019 to 2023. Coefficients ranged from 0.001 to 0.79 at 5% significance. Thirty-two districts demonstrated consistent significant correlations across all three periods, concentrated in the north and northwestern regions, where rainfall is more stable.

#### Relative humidity

Relative humidity showed the strongest and most spatially consistent associations, outperforming rainfall and temperature (Fig 4b). Correlations greater than or equal to 0.3 were observed in 60 districts across all three time periods, with coefficients above 0.9 in Serenje and Shiwang’andu. This indicates that humidity is a dominant predictor of year-round malaria suitability, supporting mosquito survival and parasite development even outside the peak rainy months.

**Fig 4b:**
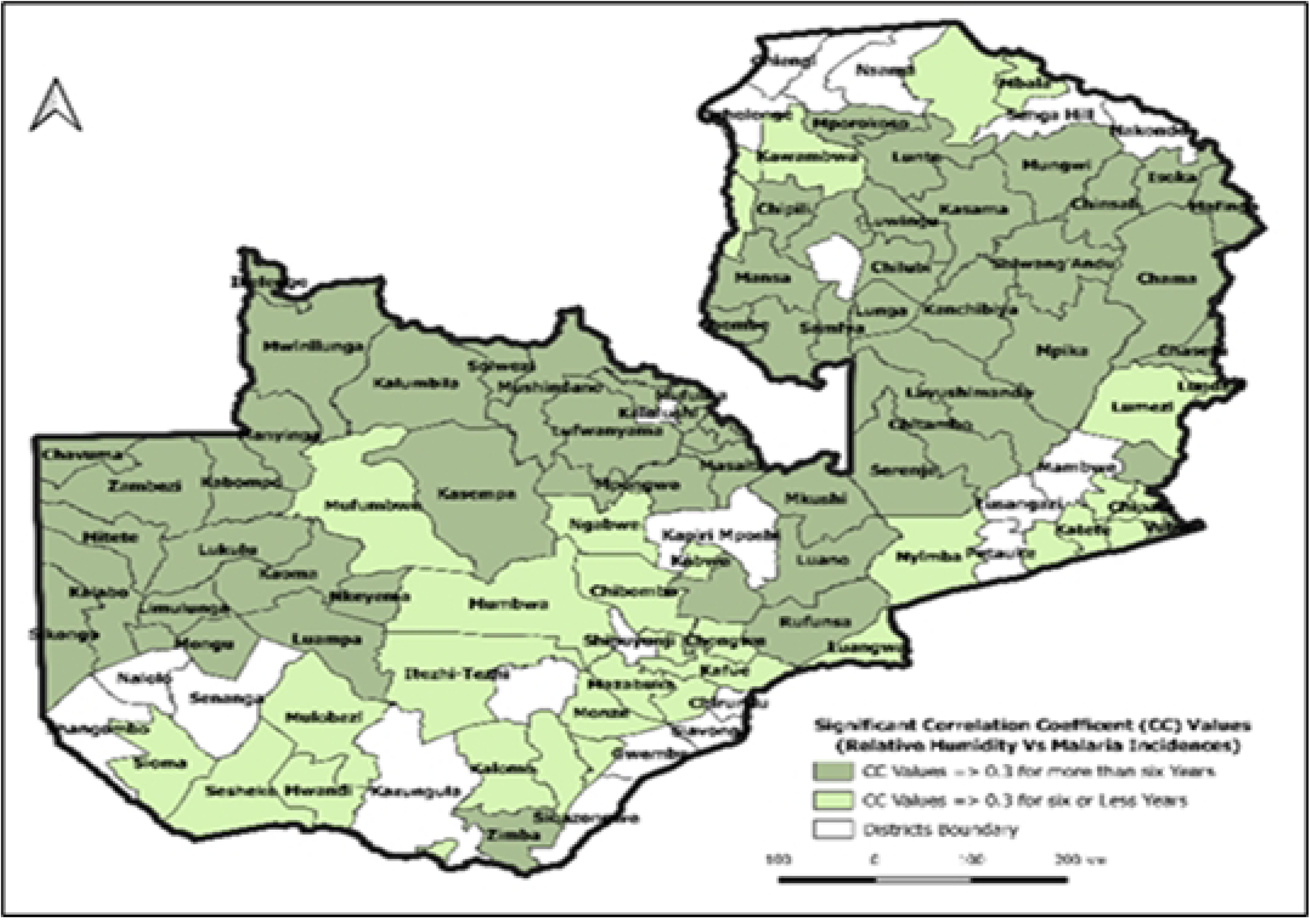
District correlation between relative humidity and malaria incidence (2009-2023). Relative humidity showed stronger correlation in 80, 56, and 82 districts with coefficients ≥ 0.9 in Serenje and Shiwang’andu. Sixty districts maintained a correlation > 0.3 across all three timeframes, suggesting that humidity plays the dominant role in sustaining malaria transmission across north and central Zambia.

#### Temperature

Temperature correlations were more localised and weaker overall. The highest association (r = 0.661) was observed in Chama District (Eastern Province). Sixteen districts showed significant correlations across all timeframes, and 32 showed significant correlations in at least two (Fig 5). These associations were mainly in valley regions spanning the Eastern, Muchinga and Southern Provinces, areas that maintain stable, warmer temperatures conducive to mosquito and parasite development.

**Fig 5a:**
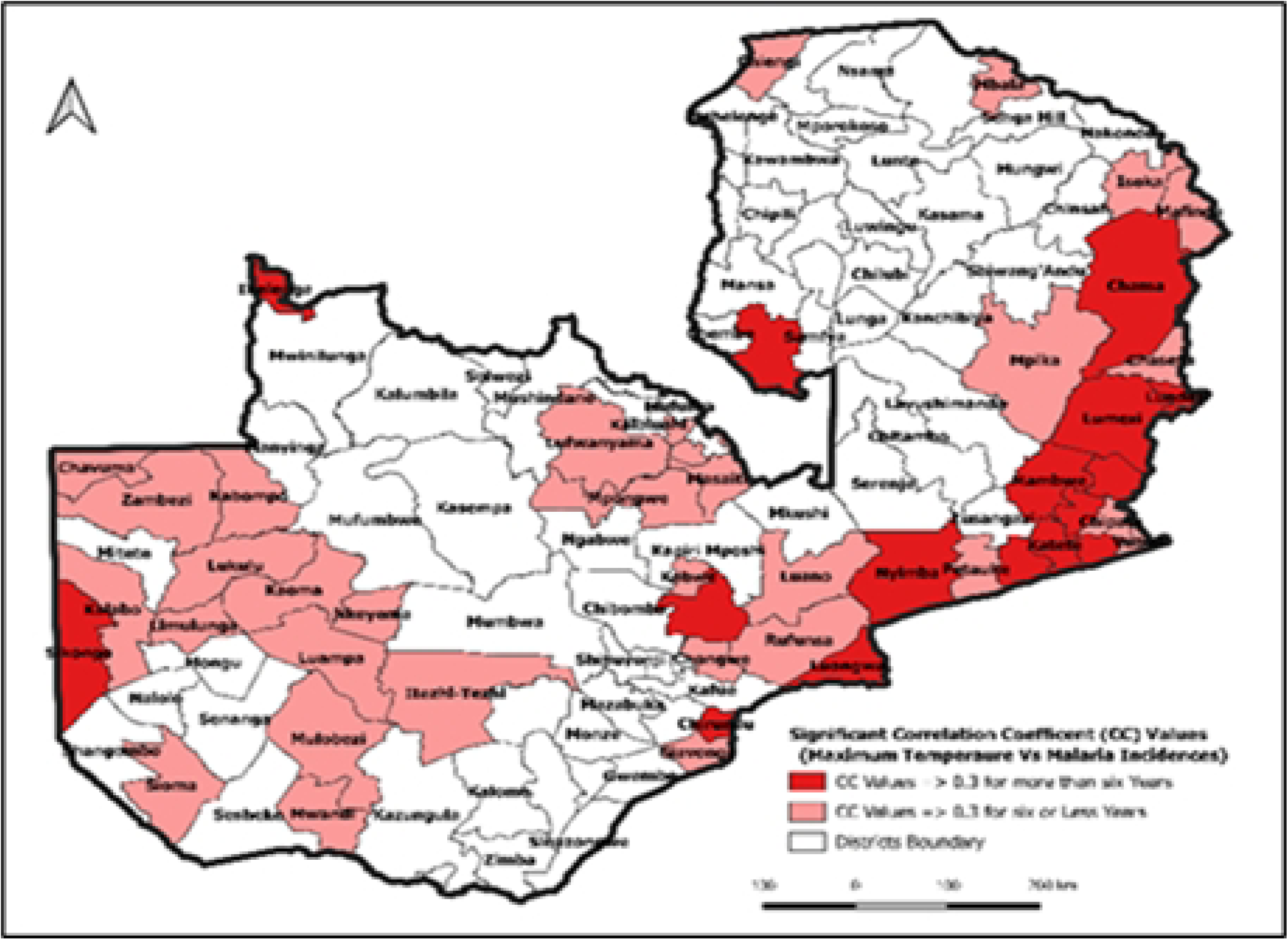
District correlation between maximum temperature vs malaria incidence.

**Fig 5b:**
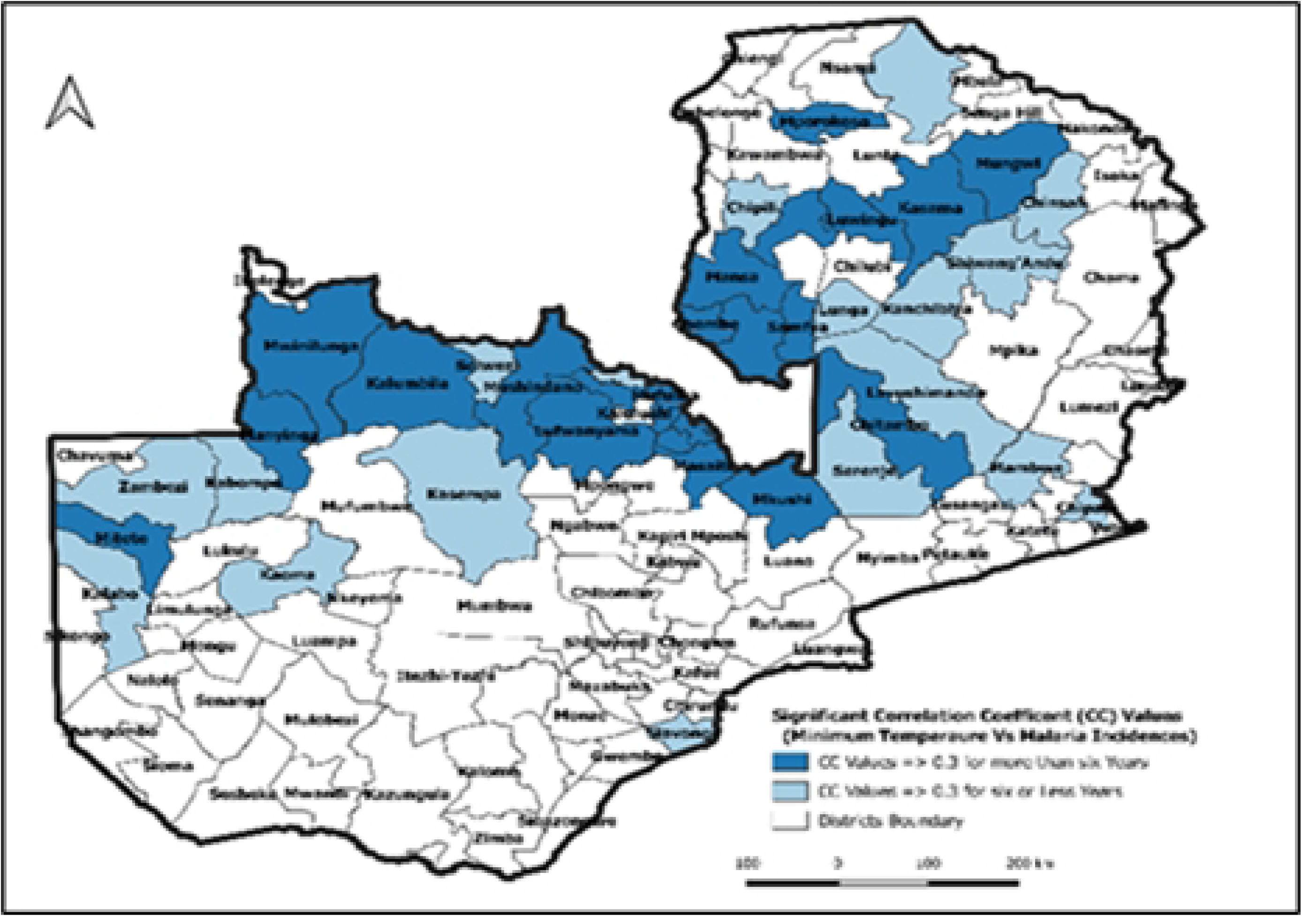
Districts’ correlation between minimum temperature and malaria incidence. The spatial distribution highlights localised but significant temperature–malaria associations in Zambia’s low-lying valleys, underscoring temperature’s secondary but facilitative role within optimal ecological zones.

### Composite Climate Correlations

A composite analysis identified nine districts, Kalabo, Kaoma, Zambezi, Kabompo, Kitwe, Luanshya, Lufwanyama, Masaiti, and Milenge, that showed correlation coefficients above 0.3 with at least two climatic parameters across multiple time frames (Fig 6). These districts demonstrated overlapping influences of rainfall and temperature variables. Additionally, a broader band of 70 districts in northern Zambia showed significant correlations between relative humidity and rainfall (Fig 6b). This highlights a large zone of sustained climatic suitability for malaria transmission driven primarily by moisture-related variables.

**Fig 6.**
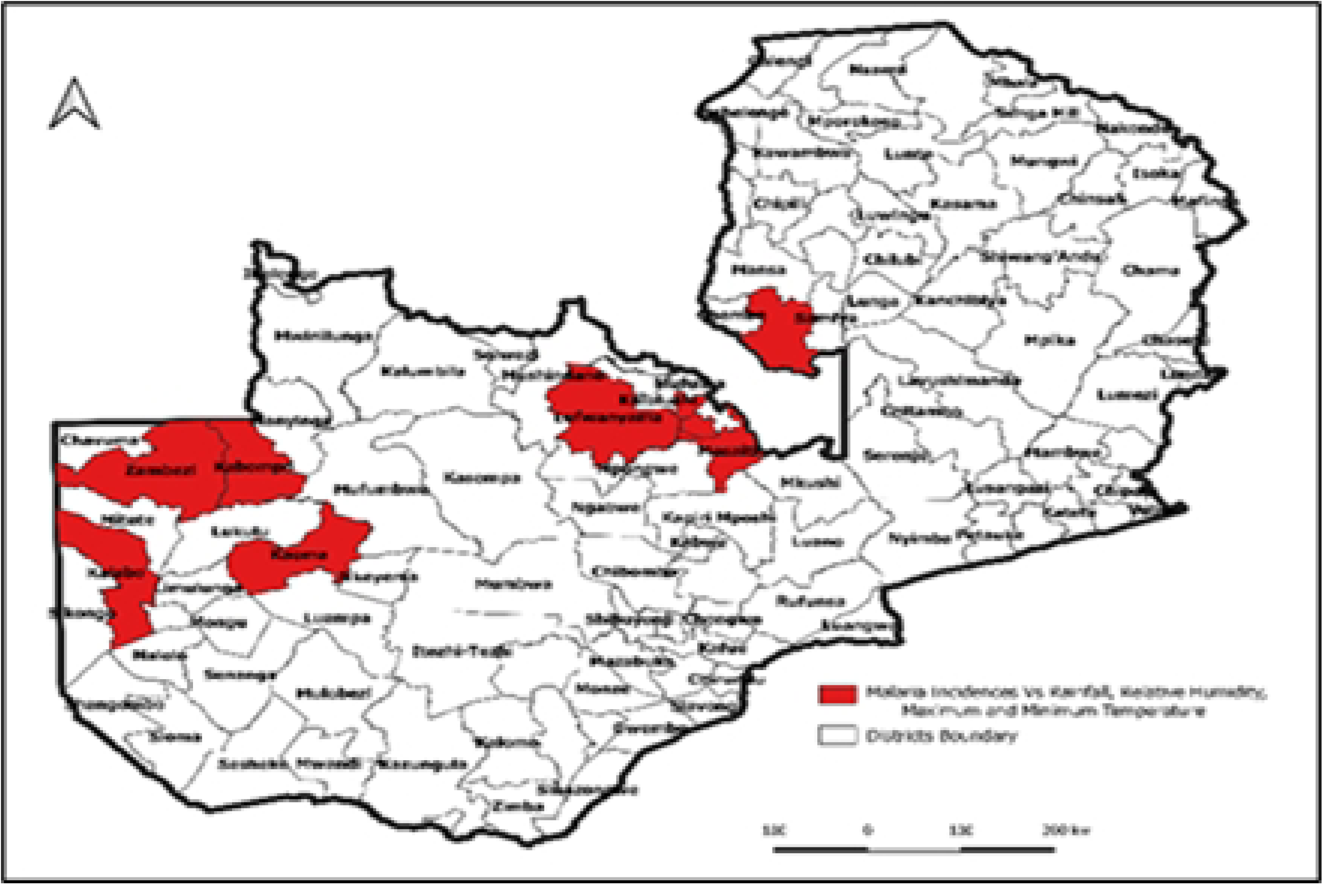
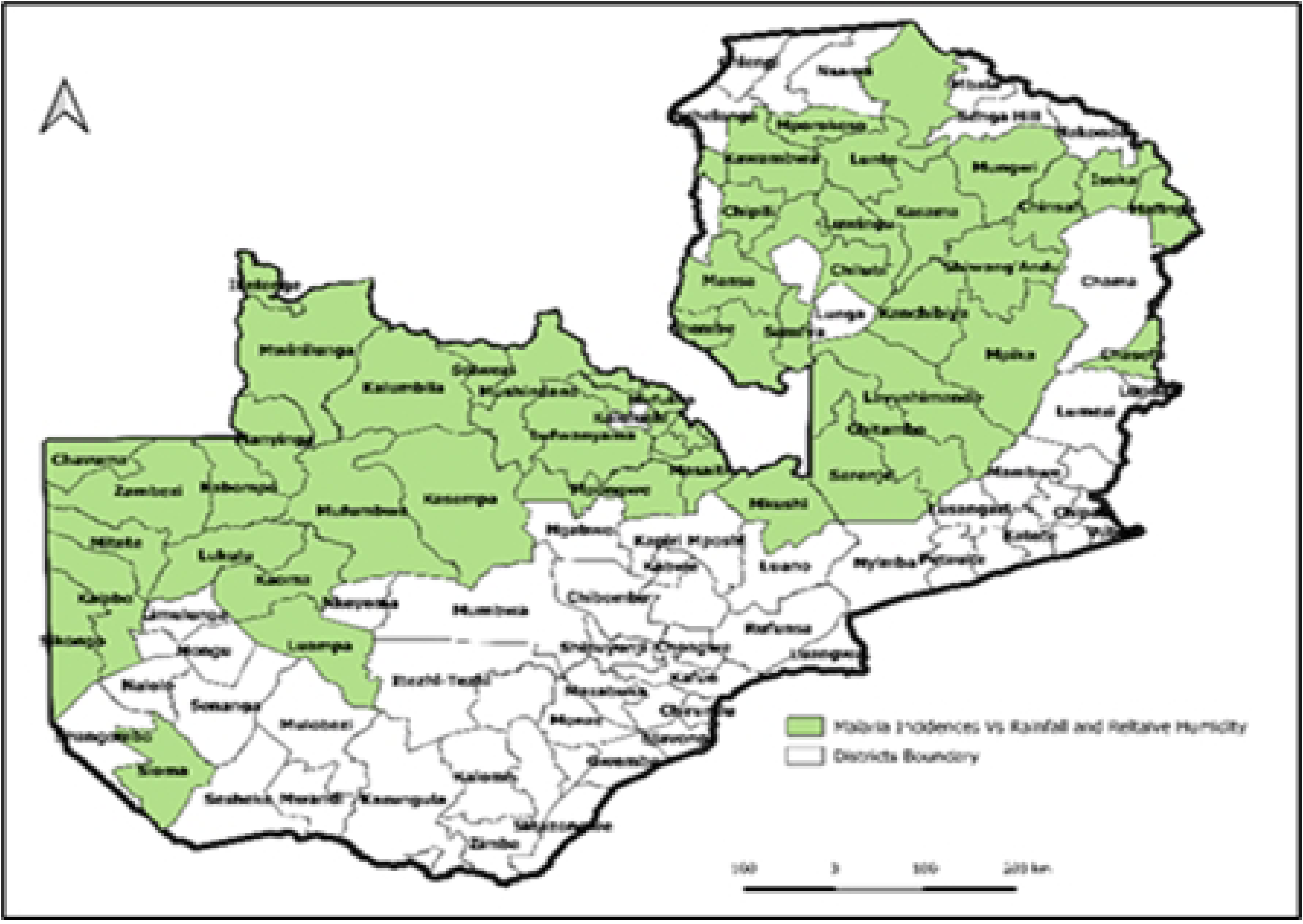
Climatic drivers of malaria incidence across Zambia (2009–2013) **(a) District-level correlations between malaria incidence and key climatic variables,** including temperature, rainfall, and relative humidity, from 2009–2013. Shading intensity represents the strength and direction of the correlation, with darker colours indicating stronger associations. This panel highlights spatial heterogeneity in climate–malaria relationships during the first five-year analysis period. **(b) Districts showing combined effects of relative humidity and rainfall on malaria incidence,** illustrating areas where both variables jointly exhibited significant positive or negative correlations with malaria trends. This panel demonstrates the spatial clustering of climate-sensitive districts and underscores the dominant role of humidity as a predictor of transmission.

### Causality and Predictive Modelling of Malaria Incidence

The Forecast Error Variance Decomposition (FEVD) analysis showed that past malaria cases accounted for 74.9% of future variability (Fig 7). Temperature, relative humidity, and rainfall explained 12.8%, 29.3%, and 13.3%, respectively. Humidity remains the most influential climatic predictor, consistent with its strong correlations.

**Fig 7:**
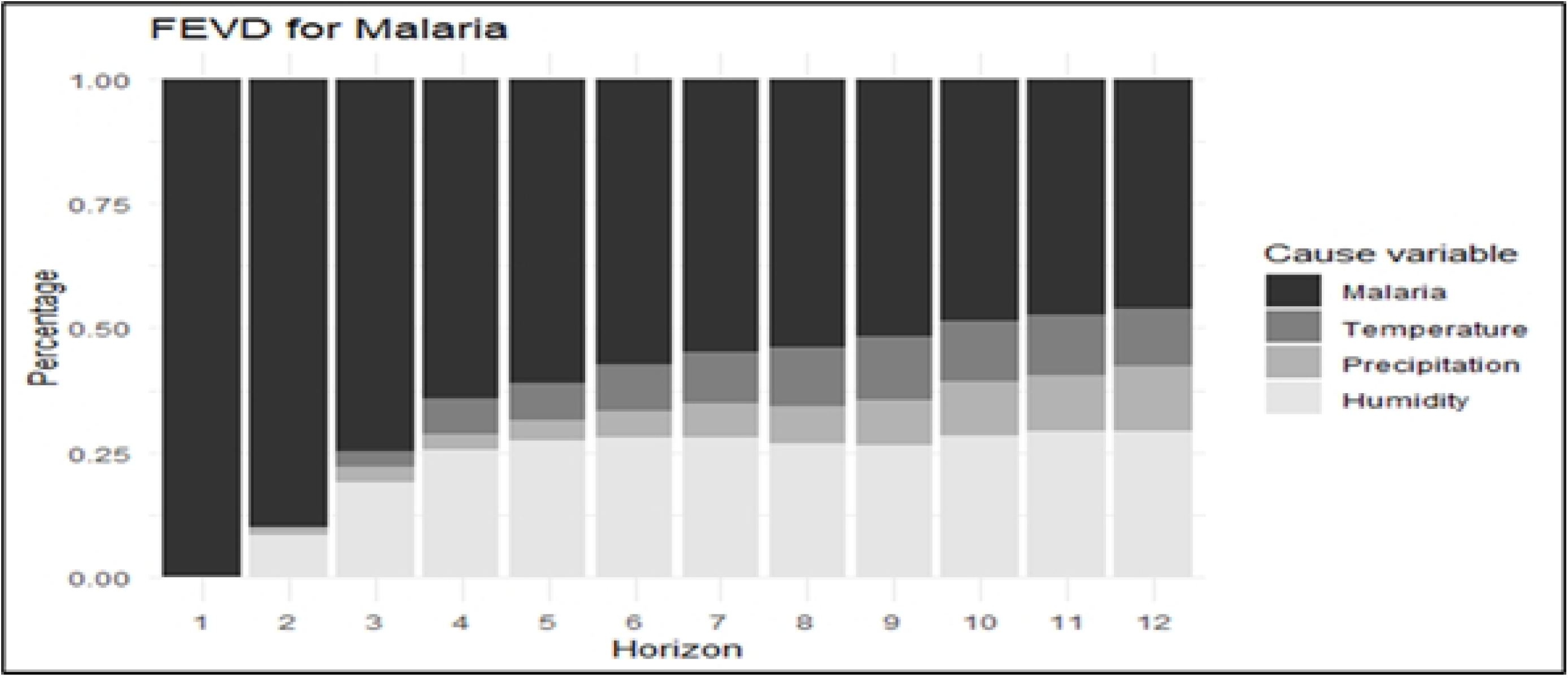
Forecast Error Variance Decomposition (FEVD) for Malaria Incidence Rates (2009-2023). Past malaria trends account for most of the model variance, but temperature and humidity also contribute significantly to interannual fluctuations, consistent with their biological roles in malaria transmission.

Impulse response analysis showed that temperature shocks produced the strongest malaria response around October, coinciding with pre-rainy season warming, while rainfall shocks produced their greatest effect around June, and humidity shocks peaked around December, respectively (Fig 8).

**Fig 8:**
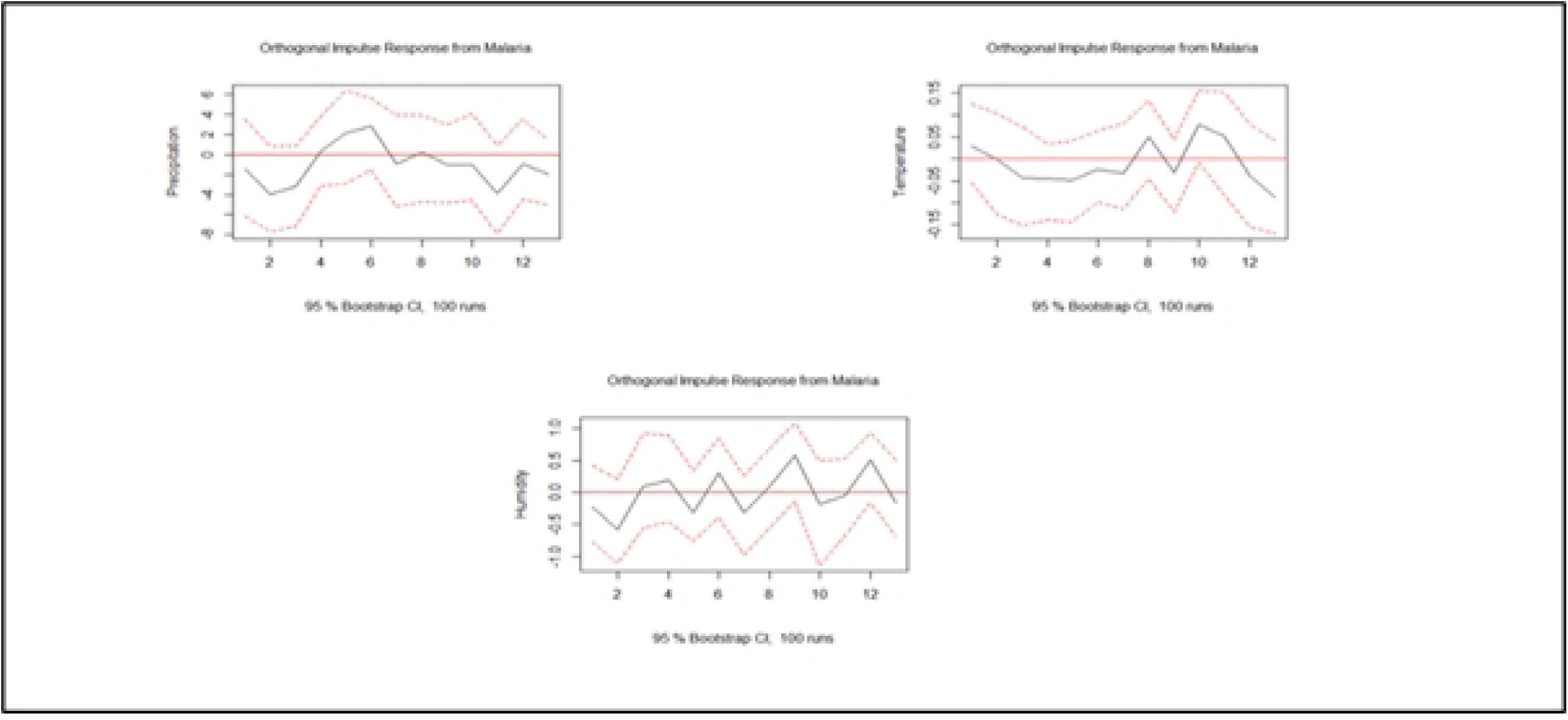
Orthogonal Impulse Response of malaria to precipitation, temperature and humidity. The dynamic response illustrates temporal lags between climatic variation and malaria transmission, with temperature and humidity exerting delayed but reinforcing effects on incidence.

Finally, predictive modelling suggests that under current trends, malaria cases will continue rising between 2024 and 2030 (Fig 9), driven by persistent climatic variability even with ongoing interventions.

**Fig 9:**
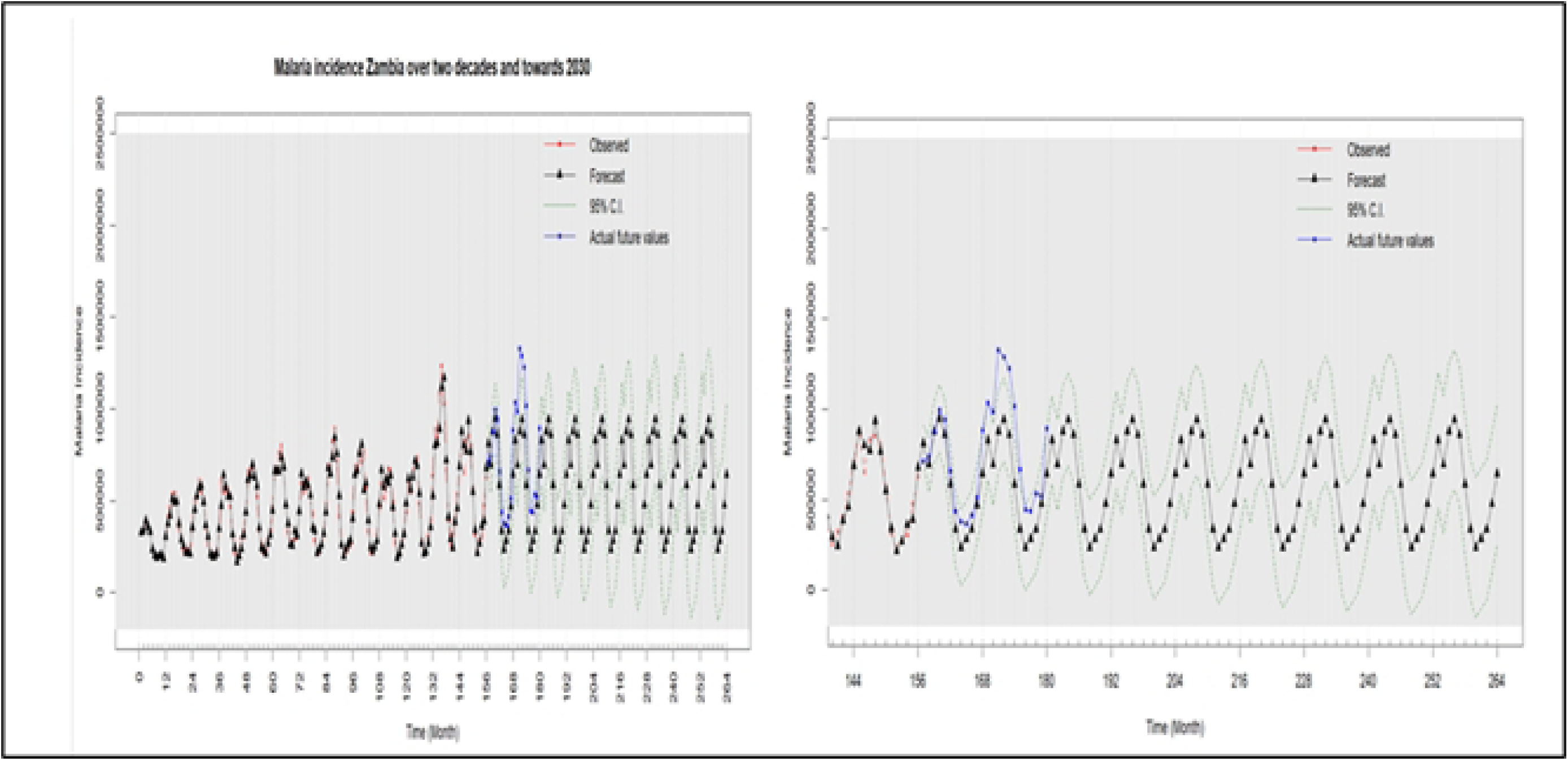
Forecasted malaria incidence in Zambia from 2024 to 2030. Model projections indicate an upward trend in malaria cases through 2030, highlighting the influence of increasing temperatures and climatic variability despite routine control measures

## Discussion

The study provides new national-level evidence linking long-term climate variability to malaria transmission dynamics in Zambia. Over 15 years of surveillance, malaria incidence showed strong seasonal alignment with rainfall and humidity, with both variables consistently preceding increases in transmission. Temperature emerged as a more localised but important driver, particularly in valley and low-lying districts, indicating that multiple interacting factors shape climatic suitability for malaria. By integrating temporal, spatial, and causality analyses, the study demonstrated that climate influences on malaria are both time-sensitive and geographically specific, underscoring the need for climate-informed elimination strategies.

Contrary to anticipated declines under Zambia’s Malaria Elimination Strategy, malaria incidence has not declined steadily. Instead, peaks have become more pronounced, and overall incidence has increased toward 2023 compared to 2009. This reversal suggests a complex interplay between climatic and non-climatic pressures, including insecticide resistance and possible antimalarial resistance, rapid population growth, urbanisation, and changes in rainfall and humidity that sustain mosquito breeding. Health system disruptions, particularly during the COVID-19 pandemic, may also have undermined consistent delivery of interventions. Together, these findings underscore the urgency of investing in innovative vector control tools, strengthening climate-informed surveillance and enhancing programmatic adaptability to protect gains made over the last decade.

Among all climatic indicators, relative humidity emerged as the most consistent and strongest predictor of malaria incidence across regions and time frames. Predictive modelling suggests that humidity and minimum temperature, rather than rainfall alone, will shape future trends, with projected increases in malaria cases through 2030 even under current intervention levels. The persistence of transmission during Zambia’s severe 2024 drought supports the finding that, even under historically low rainfall, elevated temperatures and humidity maintained ecological suitability for malaria vectors.

The geographic distribution of malaria risk is also evolving. Historically high-burden areas in Luapula, Northern, Muchinga, and Eastern provinces remain deeply affected, while new hotspots have emerged in Northwestern, Copperbelt, and Western provinces. These expanding patterns correspond with changes in rainfall distribution, extended humidity peaks and extreme weather events (7,14). Granger causality and variance decomposition further confirm that climate variability is a key driver of both spatial shifts and seasonal extension, emphasising the need to move beyond rainfall-based early warning systems towards multi-indicator climate surveillance (19,24,25).

### Implications for Malaria Control and Elimination Strategies

The findings have direct implications for Zambia’s goal of malaria elimination by 2026. The observed extension of the transmission season from December to June suggests that current interventions, such as indoor residual spraying (IRS), distribution of long-lasting insecticide-treated nets (LLIN), and seasonal malaria chemoprevention (SMC), may need to begin earlier, last longer, or be stratified geographically. Districts showing persistent climate–malaria correlations provide a strong evidence base for adaptive subnational targeting, a strategy proven effective in other endemic settings like Burkina Faso (26) and Madagascar (27).

Southern Province provides an example of how low humidity, coupled with sustained intervention coverage, maintains a low incidence (14,28). However, this is contrasted in districts such as Nchelenge, where continuously favourable climatic conditions and operational gaps limit progress (29). These comparisons emphasise that climate-driven risk must be considered alongside program performance.

Emerging tools, particularly the RTS, S and R21 malaria vaccines, offer opportunities to build climate resilience into malaria programming. Seasonal vaccination, with or without SMC, reduces clinical malaria, severe admissions, and deaths, especially in the first year (30,31). Deploying vaccines in districts with prolonged or shifting risk periods and aligning vaccination schedules with climate-informed seasonal peaks could substantially improve impact (25,27,31,32). Integrating climate surveillance with health data will be essential for optimising vaccine timing (27), especially where humidity-driven extensions in seasonal transmission are observed.

### Strengths and Limitations

Strengths of this study include the use of a long-term national dataset, integration of multiple climate data sources validated against ground stations, and the application of complementary analytical methods. The combination of hotspot detection, semiparametric models, causality testing and variance decomposition provided a robust triangulation of results. Spatial analysis across district shapefile versions allowed accurate mapping despite boundary changes.

Limitations include incomplete rainfall data for 2024, reliance on validated satellite data to supplement missing meteorological observations and the inability to model the effects of discrete climate extremes such as flash floods and heat waves.

## Conclusions and Recommendations

Relative humidity is a stronger and more consistent predictor of malaria incidence in Zambia than rainfall alone. Minimum temperature shifts also play a meaningful role, particularly in valley districts. Malaria transmission is changing in both geography and seasonal timing, making climate variability a major obstacle to achieving elimination targets. Integrating climate adaptation into malaria elimination strategies through real-time surveillance, adaptive geographical targeting, and the strategic use of the malaria vaccine could help stabilise transmission.

Future research should assess whether trends in mosquito survivability parallel those observed in other climate-sensitive infectious diseases, and should test adaptive interventions such as environmental management, housing improvements and trash exposure indexing to reduce ecological suitability for malaria transmission (33–35).

## Abbreviations

CDT: Climate Data Tool
ETCCDI: Expert Team on Climate Change Detection and Indices
FPE: Final Prediction Error Test
FEVD: Forecast Error Variance Decomposition
IRS: Indoor Residual Spraying
ISDR: Integrated Disease Surveillance and Response
LLIN: Long Lasting Insecticide-treated Nets
NMEC: National Malaria Control Centre
SMC: Seasonal Malaria Chemoprophylaxis
VAR: Vector Associated Regressions
WHO: World Health Organization
ZNPHI: Zambia National Public Health Institute
ZamSTAT: Zambia Statistics Agency

## Acknowledgements

This work is part of ongoing efforts from the Zambia National Public Health Institute and the National Malaria Elimination Centre to understand better bottlenecks towards achieving Malaria Elimination as a Country and in the Region. Many thanks to various team members at national and subnational levels, and partners who were directly and indirectly involved in this work.

## Data availability statement

All relevant data are included in the paper or its Supplementary Information.

## Author Contributions

**Conceptualisation:** Therese Shema Nzayisenga, Nyuma Mbewe, Kelvin Mwangilwa and Nathan Kapata

**Methodology and Data Curation**: Therese Shema Nzayisenga, Nyuma Mbewe, Kelvin Mwangilwa, Nathan Kapata, Stephen Bwalya, Ignatius Banda, Cheepa Habeenzeu, Malambo Mutila, Paul Msanzya Zulu, and Loveness Nikisi

**Supervision and Validation:** Nathan Kapata, Nyuma Mbewe, Kelvin Mwangilwa,

**Writing and reviewing first draft:** Therese Shema Nzayisenga, Nyuma Mbewe, Kelvin Mwangilwa, Nathan Kapata

**Writing, reviewing and editing final version:** Nyuma Mbewe, Therese Shema Nzayisenga, Kelvin Mwangilwa, Jonathan Mwanza, Stephen Bwalya, Ignatius Banda, Cheepa Habeenzeu, Malambo Mutila, Paul Msanzya Zulu, Loveness Nikisi, Freddie Masainga, Allan Mayaba Mwiinde, Peter J. Chipimo, Nathan Kapata

## Supporting Information

**S1 Table 2: Districts with top 10 highest and top 10 lowest malaria risk.** Summary statistics for the districts’ facilities with the top ten and least 10 relative risks of annual malaria risk, maximum temperature, minimum temperature, precipitation and humidity.

**S1 Table 1: Estimating Vector Autoregressions (VARs) Using Optimal Lag Length Selection** This table presents the results of optimal lag length selection for the VAR models used to evaluate temporal relationships between malaria incidence and climatic variables (temperature, rainfall, and relative humidity). Lag length was determined using standard information criteria, including the Akaike Information Criterion (AIC), Bayesian Information Criterion (BIC), and Hannan–Quinn Criterion (HQC). The selected lag structure informed subsequent Granger-causality testing and forecast error variance decomposition analyses.

